# Optimised methods for the targeted surveillance of extended-spectrum beta-lactamase producing *Escherichia coli* in human stool

**DOI:** 10.1101/2024.04.02.24305201

**Authors:** Sarah Gallichan, Sally Forrest, Esther Picton-Barlow, Claudia McKeown, Maria Moore, Eva Heinz, Nicholas A Feasey, Joseph M Lewis, Fabrice E Graf

## Abstract

Understanding transmission pathways of important opportunistic, drug resistant pathogens, such as extended-spectrum beta-lactamase (ESBL) producing *Escherichia coli,* is essential to implementing targeted prevention strategies to interrupt transmission and reduce the number of infections. To link transmission of ESBL-producing *E. coli* (ESBL-EC) between two sources, single nucleotide resolution of *E. coli* strains as well as *E. coli* diversity within and between samples is required. However, the microbiological methods to best track these pathogens are unclear. Here we compared different steps in the microbiological workflow to determine the impact different pre-enrichment broths, pre-enrichment incubation times, selection in pre-enrichment, selective plating, and DNA extraction methods had on recovering ESBL-EC from human stool samples, with the aim to acquire high quality DNA for sequencing and genomic epidemiology. We demonstrate that using a 4-hour pre-enrichment in Buffered Peptone Water, plating on cefotaxime supplemented MacConkey agar and extracting DNA using Lucigen MasterPure DNA Purification kit improves the recovery of ESBL-EC from human stool and produced high-quality DNA for whole genome sequencing. We conclude that our optimised workflow can be applied for single nucleotide variant analysis of an ESBL-EC from stool.

## Introduction

The spread of antimicrobial resistance (AMR) in bacterial pathogens is a global threat to public health. Of particular concern are infections caused by extended-spectrum beta-lactamase (ESBL) producing *Escherichia coli*, which is on the World Health Organisations (WHO) priority AMR pathogen list^1^. ESBL-producing *E. coli* (ESBL-EC) are resistant to aminopenicillins and 3rd-generation cephalosporin (3GC) antibiotics, which are commonly used to treat infections caused by Gram-negative bacteria. ESBL-EC infections can result in a higher mortality and longer hospital stays and may require access to reserve antibiotics such as carbapenems^2–4^.

Infection prevention is preferable to cure and will concurrently reduce antibiotic use. To prevent the acquisition of enteric pathogens in care facilities, it is critical to understand their transmission pathways. This is challenging for near ubiquitous organisms such as *E. coli*, which are frequently part of a healthy gut flora and can also be opportunistic pathogens^5^. Typically, human faecal samples have been shown to contain one to three different *E. coli* genotypes at any one time^6^ although as many as nine or more different *E. coli* genotypes per stool sample have been described^7^. The number of multidrug resistant *E. coli* strains detected has also been shown to increase with host age^8^. Therefore, differentiating intraspecies diversity of different *E. coli* strains requires comparisons at single nucleotide variation (SNV) resolution^9^. To track transmission events of ESBL-EC in care settings, microbiological methods need to capture the within-host diversity of ESBL-EC within and between different samples accurately and cost-effectively, even when present at low levels. Consensus on which microbiological methods are optimal and a comprehensive comparison of the impact of different microbiological methods are currently lacking.

Currently, the most common healthcare diagnostic method of ESBL-EC isolation from human stool is direct plating on cefotaxime-supplemented MacConkey or chromogenic agar and confirming the production of ESBL using the double disk diffusion method^10,11^. This is often followed by molecular investigation of ESBL resistance genes via multiplex PCR^12,13^. While these methods are effective in identifying the presence of an ESBL-EC to inform treatment options, they lack the resolution to infer transmission because they do not allow us to clearly distinguish between closely related *E. coli*^14^. The distinction between two different clades within the same ST of *E. coli* can be as little as 70 nucleotides, therefore cannot be accurately distinguished using PCR methods^15^.

Surveillance studies focused on describing transmission pathways pre-enrich samples with low microbial load, i.e., from rectal swabs, to increase detection of present ESBL-EC^16,17^. Combined with whole-genome sequencing (WGS) this allows for accurate clustering of isolates at SNV resolution. However, there is little consensus between studies, or data on controlled comparisons, on the impact of pre-enrichment broth, pre-enrichment duration, selective agar, and method of DNA extraction has on sample diversity. Here, we describe an optimized approach for targeted surveillance of ESBL-EC from human stool samples at SNV resolution to inform transmission modelling with the potential to capture within-host diversity.

## Methods

### ESBL-EC growth dynamics in different pre-enrichment broths

Pre-enrichment broths (Brain-Heart Infusion, Tryptic Soy, Buffered Peptone Water and Davis Minimal) commonly used in the recovery of ESBL-EC from clinical sources were selected for comparison in this study. These pre-enrichment broths were prepared according to the manufacturer’s recommendations **(Supplementary Table 1)**. We used three reference strains encoding ESBLs: NCTC 13341 (bla_CTX-M-15_); NCTC 13476 (bla_IMP_); NCTC 13846 (bla_CTX-M-27_), with the latter also encoding plasmid-derived colistin resistance **(Supplementary Table 2)**. We further included a clinical isolate, CAB17W, encoding bla_CTX-M-15_ **(Supplementary Table 2)**. The growth of single ESBL-EC isolates in different pre-enrichment broths was determined to establish growth dynamics. Growth of the reference ESBL-EC (NCTC 13441) in four pre-enrichment broths (BPW, DM, BHI, and TS) was monitored every 10 minutes through regular optical density readings at 600 nm wavelength in the CLARIOstar plate reader (BMG Labtech, Ottenberg, Germany) for 24 h. A population logistic model was fitted to the resulting data with the Growthcurver package^18^. The recovery of *E. coli* from different pre-enrichment broths with varying incubation times was tested. *E. coli* (NCTC 13441) was inoculated (approximately 2 x 10^7^ CFU/mL) into 5 mL of four pre-enrichment broths (BPW, DM, BHI, and TS) and grown shaking at 37 °C and 220 rpm. After 2, 4, 6 and 8 hours, 1:10 serial dilutions of each pre-enrichment culture were performed and dilutions 10^-4^ to 10^-12^ were spotted onto LB agar and incubated at 37°C overnight. The following day, the number of colony forming units (CFUs) recovered from each broth was calculated.

Stool was collected from three healthy volunteers, defined as individuals not using antibiotics or experiencing any gastrointestinal problems. The stool was resuspended in a stool diluent solution (1:5 w/v) prepared according to the maltodextrin-trehalose method with the addition of 10% glycerol ^19^. The stool slurry was mixed using a magnetic stirrer for 15 minutes and allowed to sediment for 5 minutes. Supernatants from each of the three stool slurries were mixed in equal proportions and stored in cryotubes at -80 °C. Before use in experiments, the stool slurry was rapidly thawed in a water bath at 37°C for 10 minutes and vortexed thoroughly. To determine the optimal pre-enrichment broth and incubation time for the recovery of ESBL-EC from stool, a spiked stool model was developed using 1 mL of the pooled stool slurry and a 1 μL spike of the NCTC 13441 or CAB17W (approximately 1 x 10^5^ CFU/mL). The ESBL-non-producing *E. coli* strain (NCTC 12241) was used as a negative control to determine if the antibiotic supplement sufficiently inhibited growth. The spiked stool models were pre-enriched using TS and BPW with and without the addition of selection (1 μg/ mL cefotaxime) and incubated for 4 and 18 hours, shaking at 37 °C and 220 rpm. Following incubation, 1:10 serial dilutions of each pre-enrichment culture were made and dilutions (10^-4^ to 10^-7^ for the 4-hour incubation and 10^-5^ to 10^-8^ for the 18-hour incubation) were plated onto LB agar and CHROMagar in 10 μL spots and incubated at 37 °C overnight. The number of CFUs recovered from each broth was calculated.

### Recovery of ESBL-EC using different selective agars

Selective agar (MacConkey, Membrane Lactose Glucoronide Agar (MLGA) and CHROMagar) commonly used in the recovery of ESBL-EC from environmental, clinical and food sources were selected for comparison in this study. These selective agars were prepared according to the manufacturer’s recommendations **(Supplementary Table 1)**. MacConkey and MLGA agars were supplemented to a final concentration of 1 µg/mL cefotaxime (Melford, Suffolk, UK). To assess the recovery of ESBL-EC from stool using selective agar, the stool spike model described above was used. The spiked stool was pre-enriched in BPW for 4 hours, shaking at 37°C and 220 rpm. Following incubation, 1:10 serial dilutions of each spiked stool sample were made, and 10^-4^ to 10^-7^ dilutions were plated onto MacConkey agar, CHROMagar and MLGA plates in 10 μL spots and incubated at 37 °C overnight. The number of CFUs recovered from each selective agar was calculated.

### Limit of ESBL-EC detection with pre-enrichment compared with direct plating

A series of 100 μL of CAB17W ranging from 1 CFU/mL to 1 x 10^8^ CFU/mL was spiked into 100 μL of stool slurry. The spiked stools were serially diluted (1:10) and 10 μL of each dilution was spotted onto cefotaxime supplemented MacConkey agar and incubated at 37 °C overnight. A 96 deep well plate was then inoculated with 10 μL of the remaining stool spikes in 1 mL of BPW, this was done in triplicate for each spike input concentration. The deep well plate was then sealed with a breathable sealing membrane and placed in an Innova 42R 19 mm Orbit shaking incubator set at 37 °C and 220 rpm. After 4 hours the plate was removed from the incubator, each well was again diluted in series 1:10 and spots of 10μL were plated onto cefotaxime supplemented MacConkey and incubated at 37°C overnight. The number of CFUs recovered were then calculated.

### DNA extraction kit comparison

A single colony of each of four *E. coli* strains (**Supplementary Table 2:** NCTC 13441, NCTC 13476, NCTC 13846, CAB17W) were inoculated in 5 mL of LB with and grown overnight. Then, cultures were diluted to an OD600 of 2 (∼ 2 x 10^9^ CFU/mL). The cells were washed three times in phosphate-buffered saline (PBS). The resulting pellets were resuspended in 1 mL of PBS and split into three tubes with 250 µL and used as the input for each extraction method. To assess the yield, quality, efficiency, and cost of five commercially available kits **(Supplementary Table 1)** and a simple boiling method^20^, we extracted DNA from each of the four *E. coli* isolates in technical triplicates (n=12 for each extraction method), with three experimental replicates (n=36 extractions per method) on different days. The DNA yield was then measured using the Qubit 4 Fluorometer (Agilent Technologies, California, USA) and the Agilent TapeStation System 4150 (Agilent Technologies, California, USA) was used to determine the DNA integrity. The DNA quality was assessed by measuring the A260/230 and A260/280 absorbance ratios using the NanoPhotometer (Implen, California, United States). Scores **(Supplementary Table 3)** were allocated according to the criteria in **Supplementary Table 4**. Each DNA extraction method was then ranked according to the score given, from highest (1) to lowest (6) preforming in each category: DNA yield, DNA quality, cost, hands-on time, ease of protocol and total protocol time.

### Single nucleotide variant analysis

To determine whether our protocol would provide sufficient resolution for the detection of single nucleotide variants in a stool sample, we processed a spiked stool sample and an ESBL-EC positive rectal swab. For the spiked stool sample, we inoculated 10 μL of a 100-fold diluted overnight CAB17W culture (∼ 8 x 10^7^ CFU/mL) into 1 mL of the pooled stool slurry, vortexed the mixture thoroughly and added 100 μL to 5 mL of BPW in a 15 mL Falcon tube. The ESBL-EC positive rectal swab was obtained with permission from a participant in an observational cohort study of faecal ESBL carriage in hospital patients and care home residents across Liverpool, UK. The rectal swab was placed directly in 5 mL of BPW in a 15 mL Falcon tube. Falcon tubes were incubated for 4 hours at 37 °C, shaking at 220 rpm. After 4 hours, 100 μL of the culture was spread on cefotaxime supplemented (1 µg/mL) MacConkey agar and incubated at 37 °C overnight. The following day, we picked 7 single colonies, restreaked each colony on another cefotaxime supplemented MacConkey agar and incubated at 37 °C overnight. DNA was then extracted from each of the purified colonies using the MasterPure Complete DNA and RNA Purification Kit (Lucigen, Wisconsin, USA) according to the manufacturer’s recommendations. The DNA concentrations were then measured using the Qubit 4 Fluorometer (Agilent Technologies, California, USA) and the DNA sent for whole-genome sequencing using the NovaSeq platform (Illumina Inc, California, USA) at Azenta Life Sciences (Frankfurt, Germany). Sequence data (reads) were obtained as fastq files. Sequencing adaptors were removed from the reads using Trimmomatic version 0.39^21^ and the quality of the trimmed reads was assessed using the quality metrics provided by FastQC version 0.11.9^22^. ARIBA^23^ was used to call the AMR genes using the CARD database^24^ and to determine ESBL-EC multilocus sequence type as defined by the seven-gene Achtman scheme^25^ hosted at pubMLST (https://pubmlst.org/). Single nucleotide variants (SNVs) were called between the relevant reference genome (ST167: GCF_025398915.1; ST131: GCF_004358405.1; ST1193: GCF_003344465.1) and the single pick sequences using snippy version 4.3.6 (https://github.com/tseemann/snippy). We then generated a core genome alignment of all the single pick sequences using snippy-core (with default settings). All non-nucleotide characters and potentially recombinant sequence regions were removed from the multisequence alignment using the snippy-clean_full_aln and Gubbins version 2.3.4^26^ respectively. We then extracted the SNVs from the cleaned multisequence alignment using snp-sites (using the -c option) and created a pairwise SNV distance comparison between each single pick sequence using snp-dists version 0.8.2 (https://github.com/tseemann/snp-dists).

### Statistical analysis

Measurements were summarised as mean and standard deviation or 95% confidence interval, calculated as one standard deviation above and below the mean value. Box-and-whisker plots, where presented, show medians as a horizontal line, 25th and 75th centiles as boxes, and at most 1.5 times interquartile range as whiskers. Analysis of variance (ANOVA) tests were used to determine the significance of any observed differences between groups. P < 0.05 was considered statistically significant and, where appropriate, P values were adjusted for multiple comparisons using the Bonferroni correction. A post-hoc Tukey test was used for pairwise sample comparisons when ANOVA tests were significant. All statistical analysis was performed using R version 4.1.3^27^. All R code is accessible from our Github repository (https://github.com/SarahGallichan/ESBL-EC_methods.git)

## Results

### ESBL-EC recovery using different pre-enrichment conditions and selective agars

We assessed the growth dynamics of two ESBL-EC strains (NCTC 13441 and CAB17W) in different pre-enrichment broths to determine the optimal incubation time for recovery of bacteria in log growth phase **(Supplementary** Figure 1). To summarise the dynamics of the resulting growth curves, the data was fitted to a population logistic model **(Supplementary Table 5)**. ESBL-EC reached midpoint of log-phase after approximately 4 hours in BHI, TS, and DM; and after approximately 7 hours in BPW.

Therefore, we chose incubation times of between 2 and 8 hours to quantify ESBL-EC recovery from different broths and assessed bacterial count in CFU from each broth after 4 incubation time points (2, 4, 6 and 8 hours). Across all 4 time points, BHI resulted in the highest and DM the lowest *E. coli* recovery **(Supplementary** Figure 2). We performed pair-wise broth comparisons at each time point. There was no significant difference between *E. coli* growth in BHI, BPW and TS across all 4 time points **(Supplementary Table 6)**. However, *E. coli* counts in DM was significantly lower than BHI and BPW after 2 hours; BHI, BPW and TS after 4 hours; and BHI and TS after 8 hours of incubation. The largest increase in amount of *E. coli* CFU recovered between the two time points for each broth was from 2 to 4 hours of incubation **(Supplementary Table 6)**, which is within the log growth phase of *E. coli* in each broth. Therefore, we selected 4 hours of incubation to investigate the growth of ESBL-EC from spiked stool.

We selected TS (a high nutrient broth with multiple amino acid sources) and BPW (a low nutrient broth with one amino acid source) for the spike experiments, adding defined amounts of ESBL-EC (NCTC 13441 and CAB17W) to stool samples. Donated stool was confirmed to have no growth of cefotaxime (a 3GC) resistant bacteria by plating on 1µg/mL cefotaxime supplemented MacConkey. We assessed the recovery of these spiked strains, testing whether the addition of selection with cefotaxime and the length of incubation time influenced the concentration of spiked ESBL-EC strains we were able to recover **(Figure 2)**. After 4 hours of incubation, there was no significant difference in CAB17W growth (i.e. CFU concentration) between TS and BPW and whether antibiotic was supplemented or not. However, at 4 hours the growth of NCTC 13441 in the TS without antibiotic supplement was 2.75 CFU/mL (CI: 2.28 – 3.33, p = 0.0004) more than in BPW without supplement, while there was no significant difference between the growth in either broth at 4 hours when supplemented with cefotaxime.

**Figure 1.**
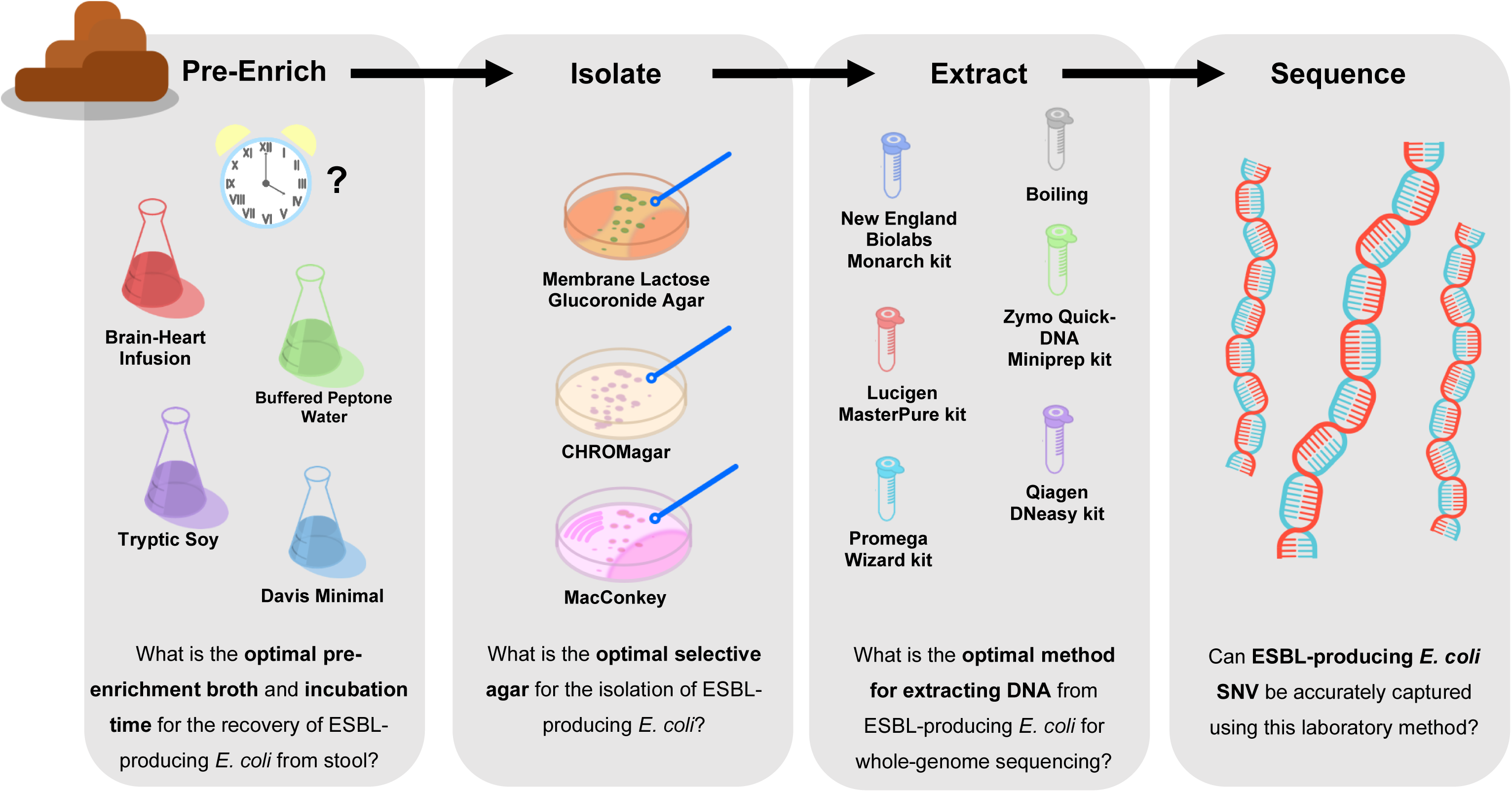
Graphical summary of the main research questions and comparisons preformed for each step of the microbiological processing of ESBL-EC from stool

**Figure 2.**
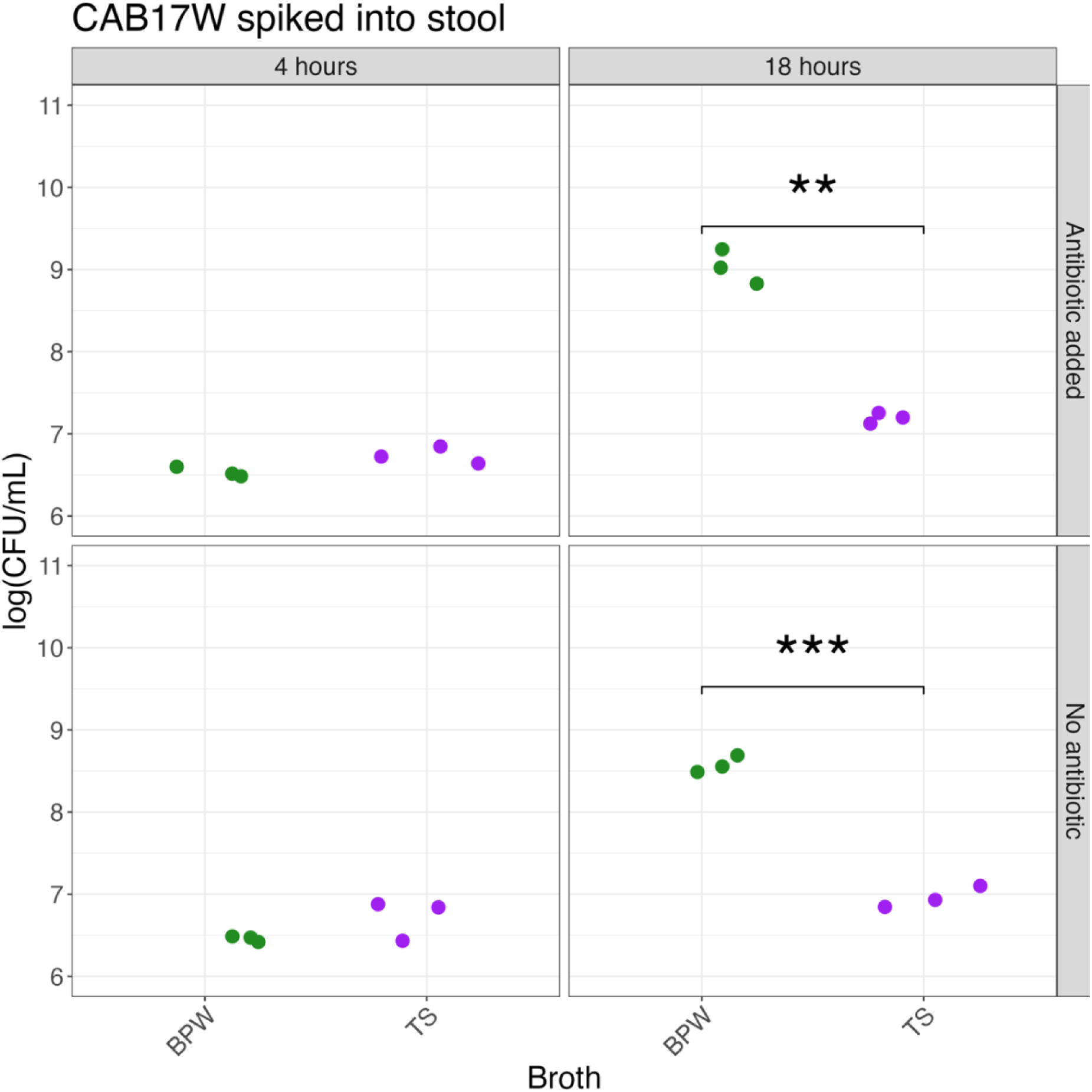
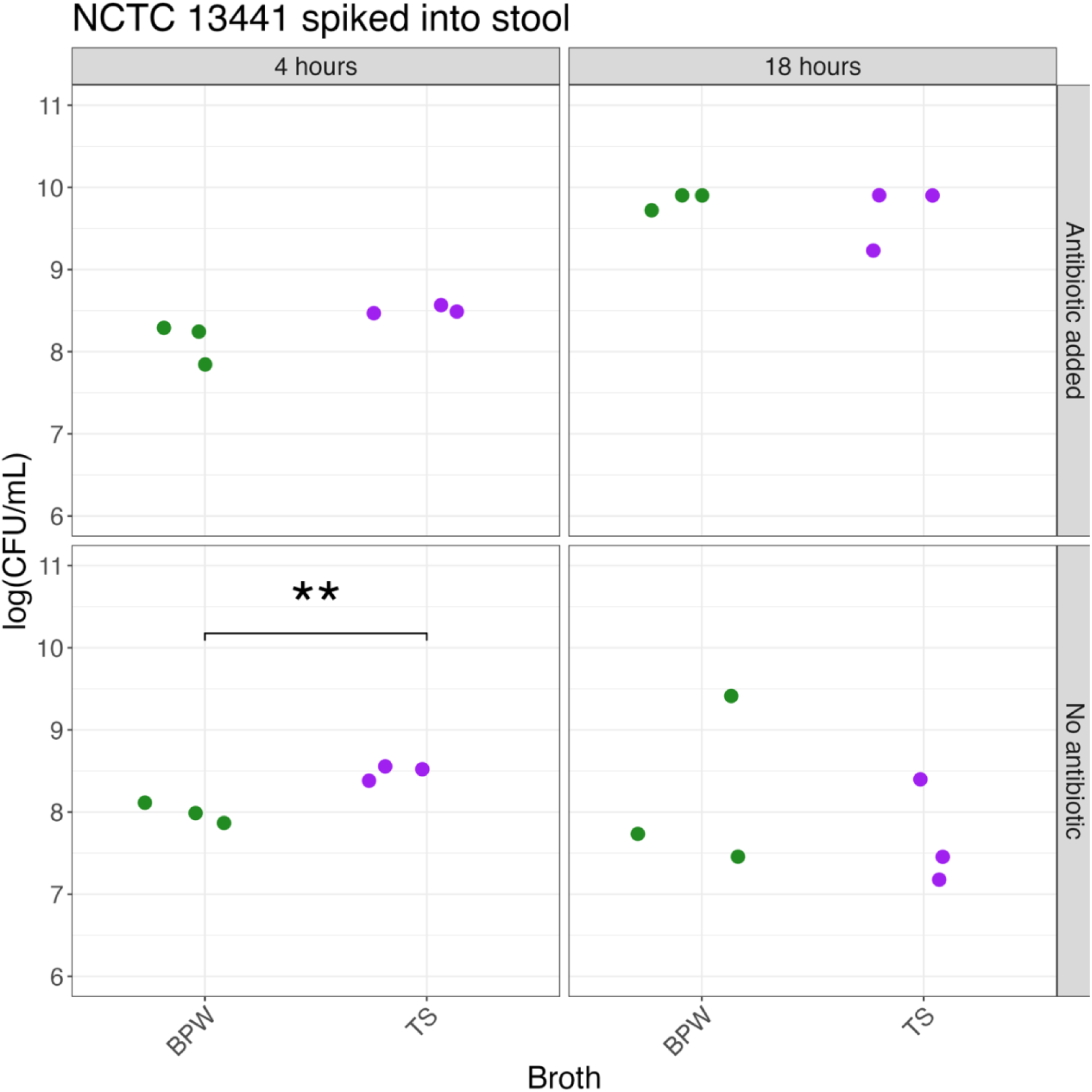
Growth of reference (NCTC 13441) and clinical (CAB17W) ESBL-EC spiked into human stool under different pre-enrichment conditions. The spiked stool models were pre-enriched in different broths (Buffered Peptone Water and Tryptic Soy) for incubation times (4-hour and 18-hour), with and without the addition of the antibiotic cefotaxime.

At 18 hours the effects of the addition of cefotaxime and broth on growth were different for each strain. Antibiotic supplement significantly increased the recovery of NCTC 13441 after 18 hours compared to no supplementation in TS, by 100.78 CFU/mL (CI: 39.81 – 251.19, p = 0.01). There was no significant difference in growth of NCTC 13441 at 18 hours between TS and BPW. However, recovery of CAB17W was significantly higher in BPW after 18 hours of incubation compared to TS, by a difference of 68 CFU/mL (CI: 51.11 – 91.26, p = 0.0001) with antibiotic supplement and 41.16 CFU/mL (CI: 33.15 – 51.10, p < 0.0001) without an antibiotic supplement. While an 18-hour pre-enrichment in antibiotic supplemented broth best recovered the highest CFU/mL for both NCTC 13441 and CAB17W, there were inconsistencies between broths depending on the strain. Therefore, we concluded that a 4-hour pre-enrichment in BPW was better for a more consistent increase in both ESBL-EC strains without requiring an antibiotic supplement.

Next, we then compared ESBL-EC recovery on three different ESBL-EC selective agars (CHROMagar, MLGA and MacConkey) from our stool spike models. ESBL-EC was recovered from all agars, with no significant differences in CFU/mL **(Supplementary Table 8)**. However, there was a clear difference in the cost per sample of each selective agar, with MacConkey being the most cost-effective and CHROMagar being the most expensive **(Supplementary Table 1)**.

The limit of ESBL-EC detection from CAB17W spiked stool was then determined for 4-hour pre-enrichment in BPW and compared with directly plating on cefotaxime supplemented MacConkey. Overall, the mean difference in ESBL-EC recovery between 4-hour pre-enrichment in BPW and direct plating was 47 CFU/mL (CI: 9.65 – 233.08, p < 0.001) **(Supplementary** Figure 3). The limit of detection using direct plating was 1.91 CFU/mL and with a 4-hour pre-enrichment, the limit of detection improved to 0.05 CFU/mL.

### Large variability in yield and quality of DNA using different extraction methods

DNA from the same input concentration of four different *E. coli* strains **(Supplementary Table 2)** was extracted using five commercially available kits **(Table 3)** and a simple boiling method. The DNA yield was measured using the Qubit fluorometer **(Figure 3)**. The boiling method resulted in the highest mean DNA yield overall for the Qubit (16.66 ng/mL, 95% CI: 16.16 – 17.16) while the DNeasy kit resulted in the lowest (3.28 ng/mL; 95% CI: 4.28 – 2.28) **(Figure 3)**.

**Figure 3.**
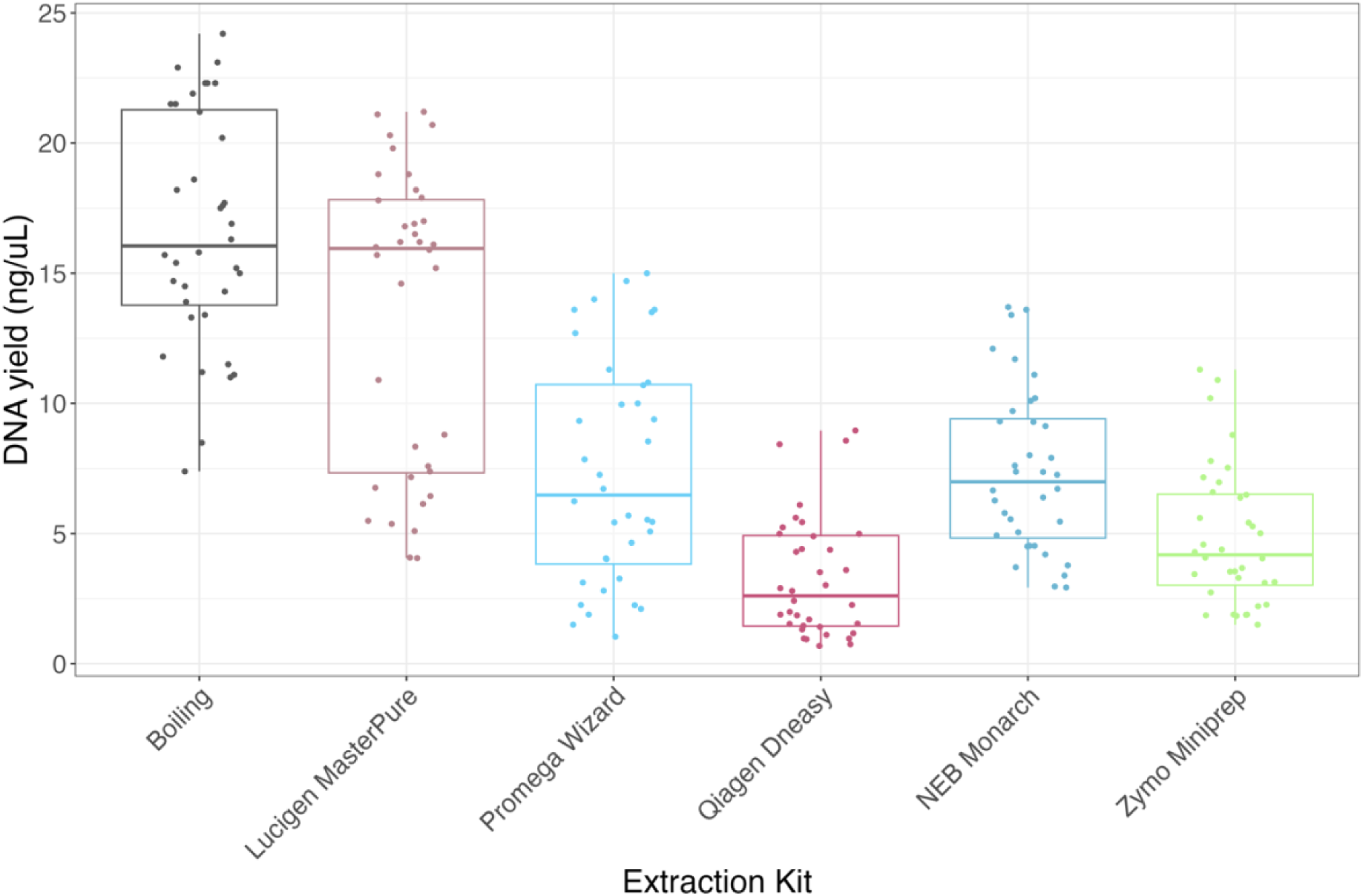
DNA yield from the six extraction methods measured using the Qubit. Technical (each sample extracted in triplicates) and experimental replicates (each extraction method performed three separate times) are plotted for each extraction kit (n = 36). The outliers are shown as points outside of the boxplot whiskers.

The DNA quality was assessed using the A260/280 absorbance ratio. Downstream applications requiring high quality DNA, such as short-read and especially long-read sequencing platforms, require that the A260/A280 absorbance ratio is in the range between 1.8 to 2.1. The Promega Wizard kit and the DNeasy kit had the largest proportion of technical and experimental replicates (67%) within this range while the Zymo Miniprep kit had the lowest proportion (11%) **(Figure 4A).** Additionally, the DNA integrity was assessed with the DNA Integrity Number (DIN), which is a numerical assessment (1 to 10) of the DNA integrity determined by the TapeStation system algorithm. A DIN score of 1 indicated very degraded DNA and DIN 10 indicated highly intact DNA. The Lucigen MasterPure and the Zymo miniprep kits had the highest DIN followed closely by the Promega Wizard **(Table 1).**

**Figure 4.**
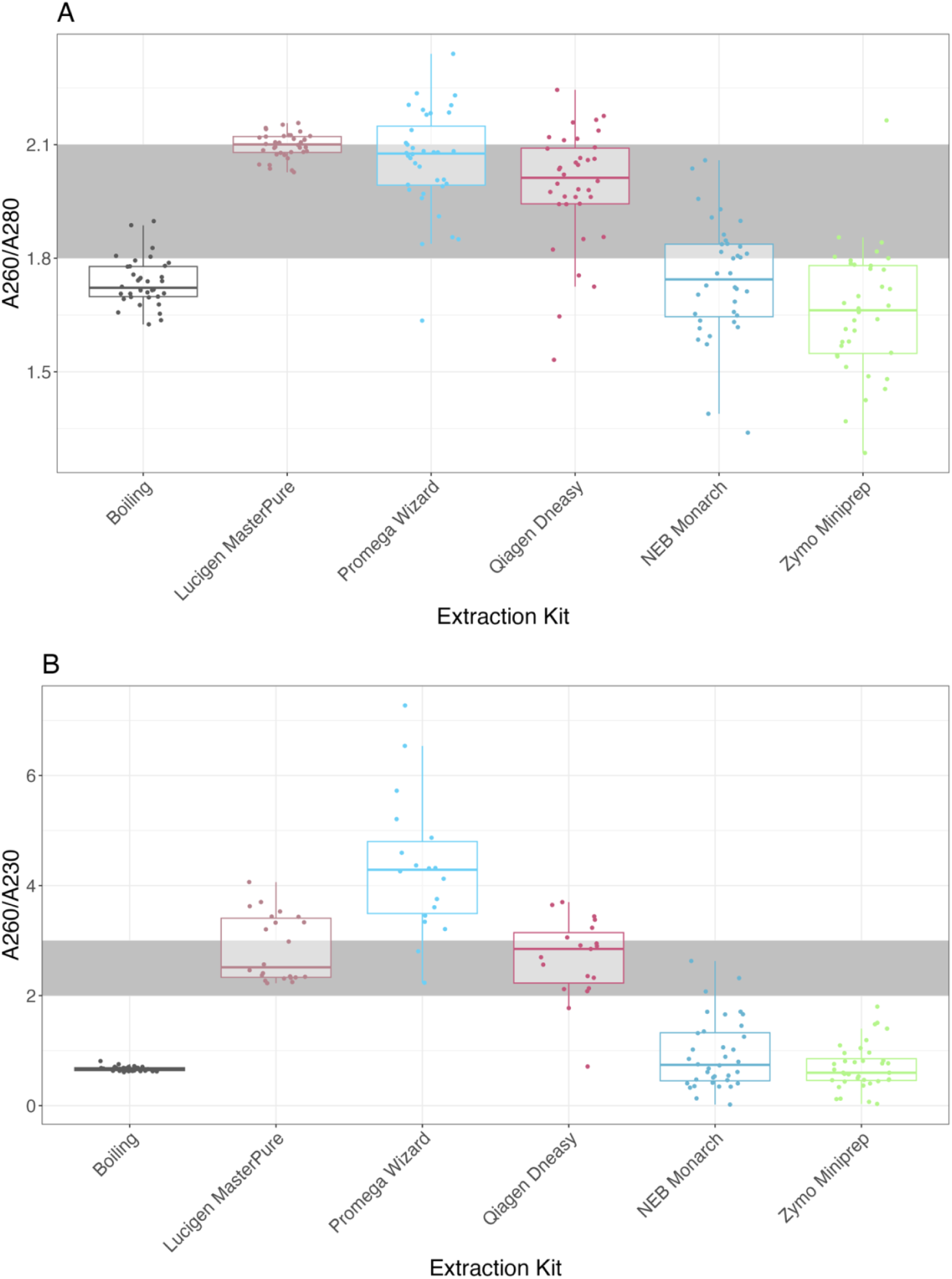
Absorbance ratios **(A)** A260/280 and **(B)** A260/230 indicating the presence of contamination and quality of DNA, respectively, of the extracted DNA extracted using six extraction methods. Technical (each sample extracted in triplicates) and experimental replicates (each extraction method preformed three separate times) are plotted for each extraction kit (n = 36). The outliers are shown as points outside of the boxplot whiskers.

**Table 1.** Mean of measurements (technical and experimental replicates averaged across three independent experiments) for each DNA extraction method, including the 95% confidence intervals, for the tested E. coli and proportion (percentage of technical replicates) within defined absorbance ratio range.

|  | Boiling | DNeasy | Lucigen<br>MasterPure | Promega<br>Wizard | NEB<br>Monarch | Zymo<br>Miniprep |
| --- | --- | --- | --- | --- | --- | --- |
| **DNA yield**<br>**(ng/ μL)** | 16.66 ± 4.66 | 3.28 ± 1.00 | 13.37 ± 0.58 | 7.37 ± 1.62 | 7.35 ± 0.97 | 4.85 ± 0.48 |
| **Absorbance ratio**<br>**(A260/A280)** | 1.74 ± 0.06 | 1.99 ± 0.15 | 2.1 ± 0.03 | 2.06 ± 0.13 | 1.74 ± 0.15 | 1.66 ± 0.17 |
| **Proportion within**<br>**range (1.8 – 2.1)** | 14% | 67% | 50% | 67% | 42% | 11% |
| **Absorbance ratio**<br>**(A260/A230)** | 0.70 ± 0.04 | 3.00 ± 0.73 | 2.89 ± 0.60 | 4.44 ± 1.27 | 1.2 ± 0.63 | 1.74 ± 0.41 |
| **Proportion within**<br>**range (2.0 – 3.0)** | 0% | 50% | 59% | 0% | 8% | 0% |
| **DNA Integrity**<br>**Number** | 1.97 ± 1.56 | 5.40 ± 0.50 | 7.40 ± 0.35 | 7.18 ± 0.45 | 6.08 ± 0.45 | 7.40 ± 0.25 |

The purity of extracted DNA was assessed using the 260/230 absorbance ratio. DNA with a 260/230 absorbance ratio of 2.0 – 3.0 are considered pure and mostly free of contaminants (such as phenols) ^28^. Lucigen Masterpure had the largest proportion of replicates (59%) within the suggested A260/A230 absorbance range, while the simple boiling method, the Zymo miniprep kit and the Promega Wizard kit failed to yield DNA within this purity range **(Figure 4B).**

Using all the measurement data, cost, ease of protocol (technical difficulty and equipment requirements), and time aspects (hands-on and total-protocol time) from the comparison of the five commercially available DNA extraction kits, a ranking matrix **(Table 2)** was produced based on the scores for each extraction method **(Supplementary Table 3)**. Simple boiling scored highly for yield, cost, and time but low for the purity scores (A260/230, A260/280 and DIN). Commercial kits performed better in this regard, with the Promega HMW and Qiagen DNeasy ranking highest for the A260/280 and the MasterPure ranking highest for the A260/230 absorbance ratio and the DIN score. All other rankings were variable between kits. The Lucigen MasterPure Complete DNA and RNA Purification Kit performed best for the extraction of high yield and quality DNA for sequencing to investigate single nucleotide variants of ESBL-EC in stool.

**Table 2.** Ranking of different DNA extraction methods, highest (1) to lowest (6).

| DNA extraction kit | Cost | Time | | Mean DNA yield (ng/ μL) | DNA quality | Difficulty |
| --- | --- | --- | --- | --- | --- | --- |
|  |  | Hands-on | Total |  |  |  |
| Qiagen DNeasy | 5 | 2 | 3 | 2 | 5 | 3 |
| Lucigen Masterpure | 2 | 4 | 5 | 1 | 1 | 6 |
| Promega HMW | 5 | 4 | 5 | 2 | 1 | 5 |
| NEB Monarch | 3 | 1 | 2 | 2 | 4 | 2 |
| Zymo Miniprep | 4 | 2 | 3 | 2 | 1 | 3 |
| Boiling | 1 | 1 | 1 | 1 | 6 | 1 |

### Validation of workflow

To investigate whether our workflow of a 4-hour pre-enrichment in BPW, plating on cefotaxime supplemented MacConkey and extraction using the MasterPure kit was sufficient for investigation of SNVs in closely related ESBL-EC strains, we performed both a control experiment where we processed an ESBL-negative stool spiked with a single clinical ESBL-EC strain (CAB17W) and assessed an ESBL-EC positive patient rectal swab using our workflow. Seven colonies from each of the cefotaxime supplemented MacConkey agar plates were then picked and sequenced. SNV analysis of the spiked stool confirmed that there were no SNVs between any of the single colony pick genomes as expected for a clonal spike **(Supplementary Table 10)**. Analysis of the ESBL-EC positive rectal swab revealed that the patient had two different ESBL-EC sequence types ST131 that had blaCTX-M-27 and ST1193 that had blaCTX-M-15. SNV analysis of each ESBL-EC sequence type showed that there were 5 SNV differences between the ST131 single picks **(Supplementary Table 11)** and 1 SNV difference between one of the ST1193 picks and the other four single picks **(Supplementary Table 12)**. We therefore concluded that our workflow was appropriate for SNV analysis of ESBL-EC from stool.

## Discussion

Although *E. coli* is the dominating species of Enterobacteriaceae in the gut of healthy human adults, *E. coli* only constitute around 0.5% -5% of the gut microbial community^29^. This makes it difficult for methods such as shotgun metagenomics to disentangle the diversity of pathogenic and non-pathogenic *E. coli* within a highly complex human gut microbiome without using costly very deep sequencing ^6,30^. On the other end of the spectrum, the analysis of single colony picks from stool samples directly plated on antibiotic supplemented agar, as is practice in many routine diagnostic laboratories, is insufficient to capture the intraspecies diversity of ESBL-EC, which can be substantial, as each colony would need to be analysed which would be extremely time-, cost- and labour-intensive^31,32^. Both methods are thus not feasible to implement as routine tool to monitor transmission of ESBL-EC in healthcare settings. Therefore, we investigated the impact of different microbiological approaches to optimise the recovery of ESBL-EC from human stool. We compared different methods at each stage of laboratory processing; pre-enrichment, isolation on selective agar, and DNA extraction, and were able to optimise a rapid and cost-effective approach for the recovery of ESBL-EC for targeted surveillance of ESBL-EC colonisation with the potential for sub-species resolution.

Numerous transmission studies have highlighted the challenge of capturing ESBL-EC variants present in small numbers in stool^33–35^. *E. coli* are only a small part of the diversity present within complex matrices and clinical samples (e.g., stool), increasing the likelihood of a test resulting in false negatives when stool is directly plated on supplemented agar^36,37^. To improve ESBL-EC detection for targeted surveillance, previous studies used pre-enrichment broth before plating on supplemented agar and were successful in recovering multiple ESBL-EC genotypes from human stool^17^. Indeed, we also showed that pre-enrichment is an effective tool to increase sensitivity for ESBL-EC detection from stool, thus improving the amount of ESBL-EC recovered on supplemented agar.

The standard practice of pre-enriching a sample for 18 to 24 hours is not conducive to rapid diagnosis and patient intervention^38,39^. A further concern with longer pre-enrichment is the increased risk of creating competition with other lineages or species and skewing the relative abundance. Further, the potential of movement of mobile genetic elements (e.g. plasmids) can bias the recovery of the original strain^40^. We found that growth of ESBL-EC was variable and strain-dependent after an 18-hour pre-enrichment with and without the addition of an antibiotic supplement, suggesting that there is indeed potential for competition bias with longer incubation times. Therefore, we investigated a short pre-enrichment and found that a 4-hour pre-enrichment yielded sufficient growth for reliable downstream identification of ESBL-EC. Hence, we concluded that 4-hour pre-enrichment was not only preferable for consistent ESBL-EC recovery but also did not necessitate the addition of an antibiotic supplement and reduced the time and cost.

There are a variety of recommended supplemented agars available for ESBL-EC detection. Cefotaxime supplemented MacConkey agar is commonly used for surveillance studies and clinical microbiology laboratories since it is readily available and the cheapest of the three agars compared here^10,11,41^. However, chromogenic agar (CHROMagar ESBL^42^) is an increasingly popular alternative to MacConkey since it is effective at distinguishing between *E. coli* and other ESBL-producing Enterobacteriaceae using colour^38,43,44^. In addition to these two agars, we also tested ESBL-EC recovery on cefotaxime supplemented Membrane Lactose Glucuronide Agar (MLGA), which is commonly used for detecting faecal contamination in water sources^45^. We found that all three agars were effective in recovering from ESBL-EC from pre-enriched stool, allowing for flexibility with selective agar choice; we hence selected cefotaxime supplemented MacConkey as the most cost-effective in our setting.

We tested a wide variety of DNA extraction methods including commercial DNA purification kits and the boiling method as the simplest and cheapest method of DNA extraction from *E. coli*^46^. Since transmission studies require high resolution typing to clearly distinguish between two *E. coli* strains, high quality DNA is required. Commercially available kits provide standardised reagents, consumables and validated DNA extraction and purification methods. Depending on the extraction protocol, specialised equipment e.g. refrigerated centrifuges, skilled personnel, and an increased amount of time and cost are needed to extract DNA^47^. Taking this into consideration, along with the cost-per-sample, quality, and yield of DNA we compared six DNA extraction methods. While the boiling method was the simplest, cheapest, least time-consuming DNA extraction method with the highest yield and thus arguably of high value for highly time-sensitive applications or in cost-restricted setting, we show that the Lucigen MasterPure produced comparable DNA yield with much higher quality scores that were consistently reproducible.

Finally, we validated our optimised ESBL-EC recovery method using a spiked stool sample and a patient rectal swab. For the spiked stool sample, all sequenced 7 colonies were identical and clonality was confirmed at single nucleotide resolution. With the patient rectal swab, we demonstrated the value of multiple colony picks through the recovery of two different ESBL-EC sequence types and the analysis of variation within each sequence type at single nucleotide resolution. The analysis of both samples at suggests that our method is suitable for studies aiming to infer transmission using SNV. While the methods we show here have focused on the recovery of ESBL-EC from stool, we suggest that these methods are also applicable or easily adaptable for the recovery other drug resistant Enterobacteriaceae. ESBL-producing *Klebsiella* and *Enterobacter* species occupy a similar niche to ESBL-EC and are all recovered from stool through direct plating on supplemented agar^10^. With the increasing prevalence of drug resistant infections worldwide, it has become crucial to understand transmission pathways to put targeted preventative measures in place to interrupt transmission.

Our study demonstrates that a 4-hour pre-enrichment in either BPW or TS is effective for the recovery of ESBL-EC from stool without the addition of a third-generation cephalosporin and cefotaxime supplemented MacConkey, chromogenic agar and MLGA are all viable options for the recovery of pre-enriched ESBL-EC. The quality and quantity of DNA using different extraction kits is variable and not all yield high-quality DNA required for whole genome sequencing. The MasterPure Complete DNA and RNA Purification Kit produced a DNA yield that was comparable to the simple boiling method, but with higher quality scores that were consistently reproducible. Altogether this optimised workflow performs well for SNV analysis of ESBL-EC from stool and can thus be applied for genomic epidemiology and transmission studies.

## Data Availability

The R scripts used to generate the figures, supplementary figures and data analyses in this manuscript are available from the GitHub repository. 
Protocols are available on Protocols.io (dx.doi.org/10.17504/protocols.io.kxygxyk3dl8j/v). 
Reads from sequenced isolates in this study are accessible in the Sequence Read Archive (SRA) using BioProject ID: PRJNA1095376.

https://github.com/SarahGallichan/ESBL-EC_methods.git.

## Ethics

The healthy volunteers gave confidential consent for the use and storage of stool samples (LSTM Research Tissue Bank RTB/2022/007). The observational cohort study of hospital patients and care home residents in facilities in Liverpool was approved by the National Research Ethics Service Greater Manchester South ethics committee (ref: 22/NW/0343). Written informed consent was obtained from participants or consultees, as appropriate.

## Supplementary materials and data availability

Supplementary Tables 1 – 12 and Supplementary Figures 1 - 3 are accessible in the supplementary material of this manuscript. The R scripts used to generate the figures, supplementary figures and data analyses in this manuscript are available from the GitHub repository https://github.com/SarahGallichan/ESBL-EC_methods.git.

Protocols are available on Protocols.io (dx.doi.org/10.17504/protocols.io.kxygxyk3dl8j/v). Reads from sequenced isolates in this study are accessible in the Sequence Read Archive (SRA) using BioProject ID: PRJNA1095376.

## Author contributions

Conceptualisation and method development was done by S.G., S.F., M.M., E.H., J.M.L., N.A.F., and F.E.G. Investigation was undertaken by S.G., S.F., E.P.-B, and C.M. Data analysis was done by S.G., J.M.L. and F.E.G. The original draft was prepared by S.G., E.H., J.M.L., N.A.F., and F.E.G and then reviewed and edited by all authors. Supervision was provided by E.H., J.M.L., N.A.F., and F.E.G.

## Acknowledgements

We would like to thank Ross Gray for the illustrations used in Figure 1 of this manuscript, Finlay Hitchman for his assistance with the pre-enrichment broth comparison work, and all the clinical staff and study participants that facilitated this study.

