## Supplementary Figures for "Optimised methods for the targeted surveillance of extended-spectrum beta-lactamase producing *Escherichia coli* in human stool"

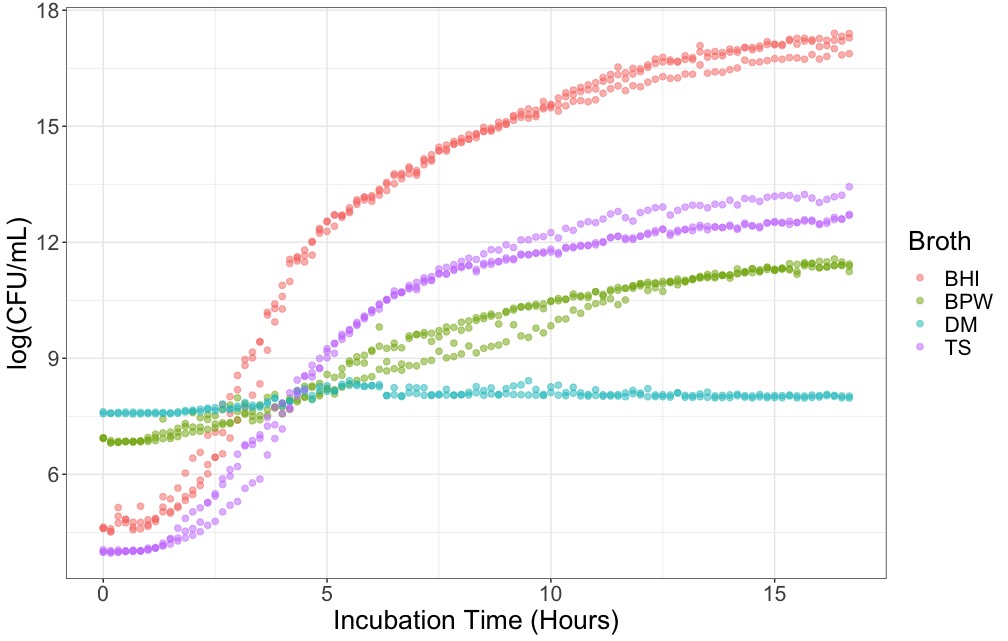

**Supplementary Figure 1** Growth of reference (NCTC 13441) ESBL-producing E. coli in different pre-enrichment broths (Brain-Heart Infusion (BHI), Buffered Peptone Water (BPW), Davis Minimal (DM) and Tryptic Soy (TS)) measured over 24 hours

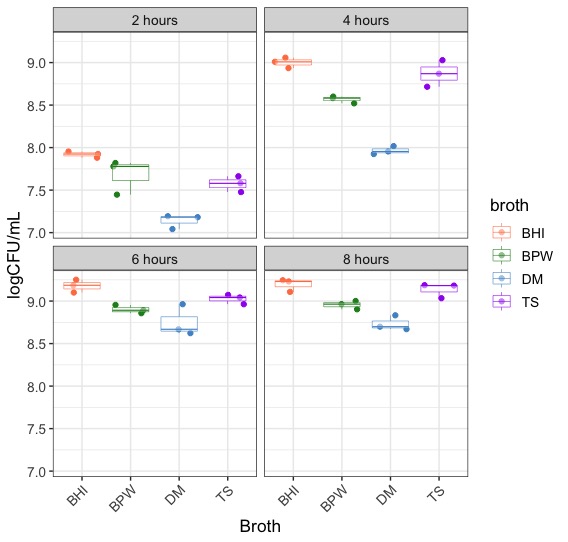

**Supplementary Figure 2** Quantitation of reference (NCTC13441) ESBL-producing E. coli after different incubation times (2, 4, 6 and 8 hours), in four different pre-enrichment broths (Brain-Heart Infusion (BHI), Buffered Peptone Water (BPW), Davis Minimal (DM) and Tryptic Soy (TS))

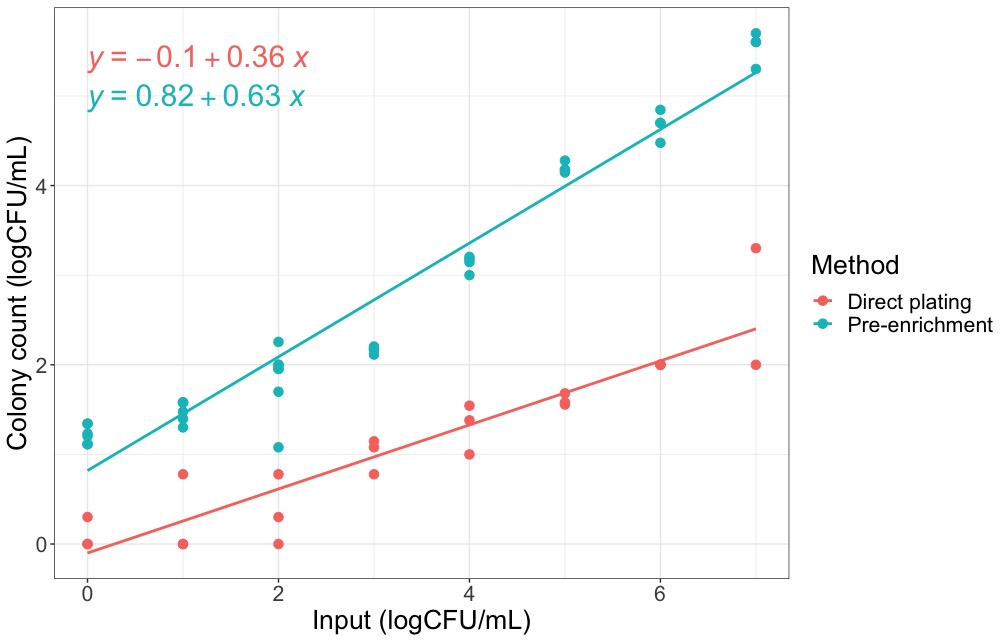

***Supplementary Figure 3*** *Colony counts for different input concentrations of a clinical ESBL-EC strain (CAB17W) spiked into stool and directly plated on cefotaxime supplemented MacConkey or pre-enriched before plating*
